# Immersive Virtual Reality Games in Neuromotor Rehabilitation with Brain-Computer Interfaces: A Scoping Review

**DOI:** 10.1101/2024.01.08.24300991

**Authors:** Mohamed N. Gaafer, Zeyad M. Ahmed, Abdullah S. El-Husseiny, Aliaa Rehan Youssef, Ahmed Al-Kabbany

## Abstract

The use of immersive virtual reality (VR) is gaining traction among the scientific community as it opens great opportunities in the field of rehabilitation. By making use of video game mechanics and Brain computer interface (BCI) that is based on electroencephalography (EEG) signals, patients undergoing neurorehabilitation can be more engaged in rehabilitation training. This paper reviews the available literature which uses BCI and VR in game rehabilitation and analyses the gaming elements and brain machine interfaces (BMI) used in each study. Four databases (IEEE Xplore, PubMed, Web of Science, Scopus) from inception until October 2023 were queried using a comprehensive searching strategy and then screened by two independent reviewers. A total of 18 articles were found eligible for qualitative synthesis. These are the main findings: (1) A diverse participant demographic spanned age and health conditions; (2) Oculus Rift has gained prominence as a VR device, replacing older CAVE systems; (3) All surveyed studies unanimously relied on the Motor Imagery (MI) paradigm, reflecting its importance in neuro motor rehabilitation and neuroplasticity; (4) Rehabilitation games displayed varied characteristics, emphasizing scoring, embodiment, and customization. Notably, the lack of gamification elements in some games suggests an area for potential enhancement and future research.

## I. Introduction

Neuromotor disabilities are widespread and vary in severity and impact on a person’s quality of life. Neurological diseases namely, stroke, cerebral palsy (CP), amyotrophic lateral sclerosis (ALS), spinal cord injury (SCI), brain injury and others result in partial or total paralysis due to neurological damage or dysfunction. Current research suggests that some diseases such as stroke and brain injury can benefit from retraining and subsequent rewiring of the brain’s neurons in a phenomenon called “Neuroplasticity”.

Conventional physiotherapy treatments have been used to attempt and induce this phenomenon, however measuring brain activity to detect patient’s intention and classify their movement to give them customised neurofeedback in what would be called a “closed loop” system, that would help guide and measure their training efforts.

Serious games, including virtual reality (VR), have been introduced in rehabilitation to improve patients’ adherence to rehabilitation protocols. Integrating gaming with BCI feedback is expected to improve patients’ adherence by allowing personalized rehabilitation, improve neuroplasticity, and train the patient to use biological signals in a more natural way. Since various body regions may be affected by neurological dysfunction, different treatment goals may be needed for rehabilitation as the ultimate rehabilitation goal is to restore whole body function, it may be beneficial to characterize VR gaming for different body region. This may help game developers in selecting modules that fulfil patient’s rehabilitation needs and integrate them in various modules.

This scoping review aims to characterise studies which have used BCI as a feedback mechanism in a closed loop neuromotor training system that makes use of immersive VR. Pinpointing the current research status in this area may help guide future research in the field.

## II. Methods

This scoping review strictly followed the Preferred Reporting Items for Systematic reviews and Meta-Analyses extension for Scoping Reviews (PRISMA-ScR) methodology.

### A. Search Strategy

Four databases (IEEE Xplore, PubMed, Web of Science, Scopus) from inception until October 2023 were searched by two independent reviewers using a combination of the following keywords. (1) BCI or Brain Computer Interface or ERP or Event-Related Potential or P300 or N200 or Motor Imagery or SSVEP or Steady state visually evoked potential or SMR or sensorimotor rhythm. (2) electroencephalography or EEG. (3) VR or Virtual Reality or XR or Extended Reality or Mixed Reality or HMD or Head Mounted Display or virtual environment. (4) Rehabilitation or training or therapy or exergame or exercise or intervention or physical therapy. (5) Motor or Paralysis or Paralyzed or neurological diseases or weakness or Muscle or Hand or Upper Limbs or Lower Limb. Relevant Boolean operators were employed. The full search strategy is given in supplementary file 1.

### B. Screening Strategy

Two independent reviewers screened the retrieved articles by title, abstract, and then by reading through the full article against eligibility criteria. To ensure accuracy and consistency, double screening was performed by two independent reviewers to confirm proper screening. Any disagreement between reviewers to resolved by consulting a third reviewers. Further, ‘sr-accelerator.com,’ which offers a range of tools, including ‘disputatron’ designed to aid in reaching a consensus and ensuring that only the most relevant and eligible articles were included in the scoping review.

### C. Eligibility Criteria

Eligible studies should include healthy individuals or those with motor impairments resulting from neurological or musculoskeletal disorders, weather adult and paediatric participants. These studies should incorporate fully immersive VR games interventions that are centred on BCI technology as a key element. No limitations are imposed on whether a comparator is present or not. Eligible outcomes may include motor function, such as strength and coordination, alongside factors related to usability, patient satisfaction, quality of life, patient compliance with the interventions, as well as adverse effects. However, the review was limited to studies published in the English language. Further, studies that do not primarily focus on motor rehabilitation or those concentrating on non-motor rehabilitation aspects were excluded. Additionally, studies not primarily centred on neuromotor rehabilitation or those employing soft exoskeletons or similar technologies are excluded from consideration.

### D. Data Charting Process

Two independent reviewers charted the following information: Reference (citation and source), Authors, Year of publication, Country of study, purpose of the study, targeted limbs for rehabilitation, BCI hardware an data acquisition including signal pre-processing techniques used, machine learning classification method applied, type of VR headset employed, any other auxiliary tools incorporated, VR scene and action characteristics, description of the reward mechanism in the virtual environment, specific diseases or conditions targeted in the study, and the number and age of enrolled participants as well as results.

## III. Results

A total of 891 documents were retrieved. After completing the whole screening process, (Figure 1) 18 were eligible for qualitative synthesis [1]–[18]. Inter-reviewer agreement was 85.11 %. Extraction was done in (Table 1).

**Table 1.** Summarized Extraction Table.

| Reference | Purpose | VR Hardware | Target Limb | VR Scene | Player Perspective | Game Reward Mechanism |
| --- | --- | --- | --- | --- | --- | --- |
| [1] | Post-stroke rehab | HTC Vive | Lower | Road with checkpoints | Riding a tricycle | Timings, scoring system |
| [2] | BCI for gait rehab | Oculus | Lower | virtual corridor | Self-avatar, First person perspective | - |
| [3] | Virtual movements agency with BCI | Oculus | Upper | Table with two buttons | Hands on table, monitor upfront | - |
| [4] | Arm rehab using MI | Oculus | Upper | Living room with beach view | Arms only visible | - |
| [5] | Upper & lower extremities rehab | FOVE | Upper & lower | Catch falling fruits, kick footballs | Full body embodiment | Progress bars, scores, explosions |
| [6] | BCI-VR in chronic stroke patients | Oculus | Upper | Personalized avatar | Set of virtual hands | - |
| [7] | MI-BCI training with embodiment | Oculus | Upper | Desk with red button | Hands laid on desk, tv upfront | Animation speed |
| [8] | BCI & VR embodiment | Oculus | Upper | - | Personal arm & target arm | - |
| [9] | MI-MO on brain activation | specialized MR-compatible video visor | Upper | Rowing boat in open sea | Rowing movement of virtual arms | - |
| [10] | VR & BCI in rehab | Oculus | Upper | Virtual arm | Arm flexion/extension | - |
| [11] | VR & MP in improving MI | Oculus | Upper | Garage door with lever | Rotating lever with hands | - |
| [12] | EEG, sEMG, & IVR in neurorehab | - | Lower | Road with checkpoints | Riding a monocycle | Scoring system |
| [13] | Use of immersive VR in MI training | Oculus | Upper | Blank space | Set of virtual hands | - |
| [14] | VR & BCI for stroke recovery | Oculus | Upper | Simple room with desk | Sitting in front of desk | - |
| [15] | VR rehab for children with CP | Oculus | Lower | Path with obstacles | Walking towards obstacles | - |
| [16] | Virtual therapist & virtual augmentation | Oculus | Upper | - | Virtual therapist's avatar | Scoring system |
| [17] | Multimodal BCI control of VR | DAVE | Lower | Ski mountain race | Penguin jumps and eats fish | Scoring system |
| [18] | VR for chronic pain assessment | NVIS SX111 | Upper | Room with mirror | Hand resting on table, sitting in front of mirror | - |

**Figure 1.**
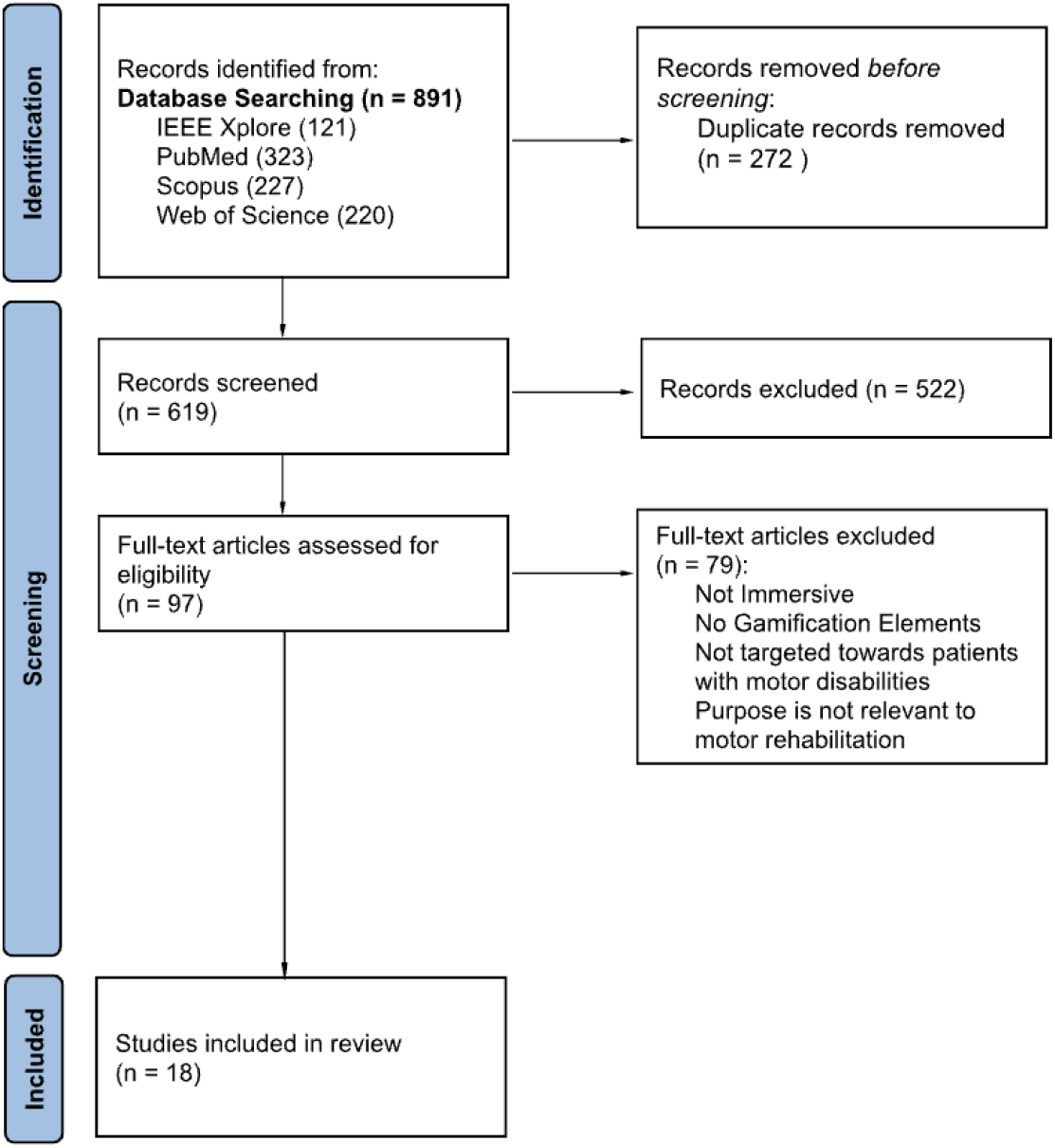
PRISMA Filtering Flowchart

The included population covered a wide age range with 5.6% focusing on children, 83.3% adult population and 5.6% elderly population (above 65). One study failed to mention participants’ age group. The majority 66.7% of studies tested the system on healthy participants only, and the largest sample size was 68. Many non-healthy participants were post-stroke or cerebral palsy patients, and suffered from paraplegia, diplegia, or tetraplegia.

Regarding the hardware used, the oculus rift was the most common VR device (61%). In studies before 2013 the CAVE system was used to achieve immersion with stereoscopic glasses. The main BCI paradigm employed was Motor Imagery (MI), all (100%) of papers relied on it either partially or totally. With some papers making use of multimodal approaches. Feedback mechanisms were used in some papers, these included Functional Electrical Stimulation (FES) and Haptic feedback.

All but one of the papers made use of a first-person avatar view, with the argument that this increases one’s embodiment. Embodiment refers to the ability of users to perceive virtual movements as their own. Some (11%) went to the extent of providing avatar appearance customization for enhanced embodiment. The virtual scenes varied between complex environments (i.e., cycling through a road, rowing a boat) to simpler and more stationary ones limited to just the movements of certain limbs.

All studies incorporated a form of visual feedback. Users were prompted with visual cues displaying the required motor function, followed by a window (∼2-10s) for reaching neural activation representing MI of said function. Successful runs are followed by the corresponding avatar animation. Only two studies mentioned a gamification system where performance was rewarded using a progress bar and a score system.

A majority of 72% of the papers reviewed targeted the rehabilitation of upper extremities, 22% targeted lower extremities, while the remaining 5% accommodated for both. A key difference between the virtual environments of upper and lower extremity rehabilitation is the continuity of MI tasks. The former provides the user with a time window for generating MI after each required task. The flow of lower extremity-targeting applications however took a more continuous approach, where constant motion (i.e., walking, cycling) through the virtual environment was observed if the required MI was sustained. In consequence, the environments created for the latter were larger and displayed a wider variety of 3D objects to enable the user to wander around with better immersion.

Outcomes revealed that immersive BCI-VR rehabilitation systems offer higher levels of performance in MI tasks when compared to non-immersive computer screen solutions. Moreover, all papers mentioning user satisfaction concluded that VR offers higher engagement and interest levels and instilled a sense of enthusiasm. A common complaint however was the onset of cybersickness, and fatigue caused by HMD/VR headsets. Three studies focusing on the effect of embodiment on neurofeedback performance all reached the same findings relating greater performance with higher embodiment levels. A minimal 16% of the papers reported direct assessments of motor/neural function improvements.

## IV. Discussion

This scoping review comprehensively examines the literature on the integration of immersive virtual reality (VR) games with Brain-Computer Interface (BCI) technology for neuromotor rehabilitation. A significant finding is the broad demographic diversity and wide age range of participants, highlighting the potential applicability of these technologies across an individual’s lifespan. While a portion of the studies includes non-healthy participants with conditions like stroke and cerebral palsy, the majority focus on healthy subjects. This underscores the need for more inclusive research involving clinical populations.

The evolution of VR hardware is also noteworthy, with the Oculus Rift becoming the predominant choice post-2016. In contrast, earlier studies tended to utilize the CAVE system alongside stereoscopic glasses to create an immersive experience. This shift reflects technological advancements and possibly user preferences and accessibility.

A universal reliance on the Motor Imagery (MI) paradigm across the reviewed studies underscores its critical role in neuromotor rehabilitation and neuroplasticity—the brain’s capacity to adapt and form new neural connections. This neuroplasticity is fostered through repetitive and intensive training, sensory feedback, motor learning, and tailoring the experience to the user’s needs. Virtual reality and serious gaming are pivotal in this context, enhancing immersion and thereby boosting engagement and motivation.

The studies demonstrate a variety of design elements within the games. Some emphasize scoring and progression to engage users, while others focus on embodiment and customization to enhance the sense of connection and personal relevance. These features collectively showcase the versatility of immersive BCI-VR games in addressing motor impairments and their potential to significantly improve rehabilitation outcomes for individuals with neurological or musculoskeletal disorders.

However, the limited focus on gamification elements in some games could potentially hinder patient engagement and motivation, indicating an area ripe for further development and research. Enhancing these aspects could lead to more effective and enjoyable rehabilitation processes, ultimately contributing to better health outcomes and quality of life for patients.

## V. Limitations

The limitations of this review include small sample sizes, with most studies involving fewer than 30 participants, and a predominance of healthy participants, limiting applicability to clinical populations. Furthermore, the focus on BCI classification and physiological outcomes over clinical efficacy narrows the scope of the findings. A notable lack of emphasis on game mechanics and reward systems, crucial for patient engagement, further limits the review. Future research should address these gaps by increasing sample sizes, including more diverse and clinical populations, conducting comprehensive clinical assessments, and enhancing game elements to improve rehabilitation effectiveness and patient motivation.

## VI. Conclusion

In conclusion, this scoping review has provided a comprehensive overview of the use of immersive virtual reality (VR) games integrated with Brain-Computer Interface (BCI) technology for neuromotor rehabilitation. The studies demonstrate a broad demographic reach and an evolving preference for advanced VR hardware like the Oculus Rift, underscoring the growing accessibility and applicability of these technologies. The universal reliance on the Motor Imagery (MI) paradigm highlights its significance in promoting neuroplasticity and enhancing rehabilitation outcomes.

However, the review also reveals critical limitations, including small sample sizes and an overrepresentation of healthy participants, which may limit the generalizability of the findings to clinical populations. The lack of focus on game mechanics and reward systems in some studies suggests a need for more engaging and patient-centric designs. Future research should aim to address these limitations by involving larger and more diverse populations, emphasizing clinical efficacy, and developing engaging game elements. By doing so, immersive BCI-VR games have the potential to transform neuromotor rehabilitation, offering more effective, engaging, and personalized therapeutic interventions.

## Supporting information

File 1 Full Search Query

## Data Availability

All data produced in the present study are available upon reasonable request to the authors

https://pastebin.com/dl/CUuPmGCL

## Acknowledgement

VRapeutic for sponsoring this research and upcoming graduation project.

ChatGPT 4 was used to assist in linguistic review of the discussion, limitations, and conclusion sections.

## Supplementary Files

File 1: Full Search Strategy

## References

[1] T. Bastos-Filho, M. A. Romero, V. Cardoso, A. Pomer, B. Longo, and D. Delisle, “A Setup for Lower-Limb Post-stroke Rehabilitation Based on Motor Imagery and Motorized Pedal,” in VIII LATIN AMERICAN CONFERENCE ON BIOMEDICAL ENGINEERING AND XLII NATIONAL CONFERENCE ON BIOMEDICAL ENGINEERING, vol. 75, no. 8th Latin American Conference on Biomedical Engineering (CLAIB) / 42nd National Conference on Biomedical Engineering (CNIB), C. A. G. Diaz, C. C. Gonzalez, E. L. Leber, H. A. Velez, N. P. Puente, D. L. Flores, A. O. Andrade, H. A. Galvan, F. Martinez, R. Garcia, C. J. Trujillo, and A. R. Mejia, Eds., Univ Fed Espirito Santo, BR-29075910 Vitoria, ES, Brazil, 2020, pp. 1125–1129. doi: 10.1007/978-3-030-30648-9_146.

[2] B. Alchalabi, J. Faubert, and D. R. Labbé, “A multi-modal modified feedback self-paced BCI to control the gait of an avatar,” J Neural Eng, vol. 18, no. 5, p. 056005, Oct. 2021, doi: 10.1088/1741-2552/abee51.

[3] B. Nierula, B. Spanlang, M. Martini, M. Borrell, V. V. Nikulin, and M. V. Sanchez-Vives, “Agency and responsibility over virtual movements controlled through different paradigms of brain−computer interface,” J Physiol, vol. 599, no. 9, pp. 2419–2434, May 2021, doi: 10.1113/JP278167.

[4] D. Achanccaray, G. Mylonas, and J. Andreu-Perez, “An Implicit Brain Computer Interface Supported by Gaze Monitoring for Virtual Therapy,” in 2019 IEEE International Conference on Systems, Man and Cybernetics (SMC), IEEE, Oct. 2019, pp. 2829–2832. doi: 10.1109/SMC.2019.8913962.

[5] T. Karácsony, J. P. Hansen, H. K. Iversen, and S. Puthusserypady, “Brain Computer Interface for Neuro-rehabilitation With Deep Learning Classification and Virtual Reality Feedback,” in Proceedings of the 10th Augmented Human International Conference 2019, New York, NY, USA: ACM, Mar. 2019, pp. 1–8. doi: 10.1145/3311823.3311864.

[6] A. Vourvopoulos et al., “Effects of a Brain-Computer Interface With Virtual Reality (VR) Neurofeedback: A Pilot Study in Chronic Stroke Patients,” Front Hum Neurosci, vol. 13, Jun. 2019, doi: 10.3389/fnhum.2019.00210.

[7] F. Škola and F. Liarokapis, “Embodied VR environment facilitates motor imagery brain–computer interface training,” Comput Graph, vol. 75, pp. 59–71, Oct. 2018, doi: 10.1016/j.cag.2018.05.024.

[8] J. M. Juliano et al., “Embodiment is related to better performance on a brain–computer interface in immersive virtual reality: A pilot study,” Sensors (Switzerland), vol. 20, no. 4, p. 1204, Feb. 2020, doi: 10.3390/s20041204.

[9] A. Vourvopoulos, D. A. Blanco-Mora, A. Aldridge, C. Jorge, P. Figueiredo, and S. B. i Badia, “Enhancing Motor-Imagery Brain-Computer Interface Training With Embodied Virtual Reality: A Pilot Study With Older Adults,” in 2022 IEEE International Conference on Metrology for Extended Reality, Artificial Intelligence and Neural Engineering (MetroXRAINE), IEEE, Oct. 2022, 10.1109/MetroXRAINE54828.2022.9967664. pp. 157–162. doi:

[10] D. Achanccaray, K. Pacheco, E. Carranza, and M. Hayashibe, “Immersive Virtual Reality Feedback in a Brain Computer Interface for Upper Limb Rehabilitation,” in 2018 IEEE International Conference on Systems, Man, and Cybernetics (SMC), IEEE, Oct. 2018, pp. 1006–1010. doi: 10.1109/SMC.2018.00179.

[11] B. Yazmir and M. Reiner, “Monitoring brain potentials to guide neurorehabilitation of tracking impairments,” in IEEE International Conference on Rehabilitation Robotics, IEEE, Jul. 2017, pp. 983–988. doi: 10.1109/ICORR.2017.8009377.

[12] V. F. Cardoso et al., “Neurorehabilitation Platform Based on EEG, sEMG and Virtual Reality Using Robotic Monocycle,” in XXVI BRAZILIAN CONGRESS ON BIOMEDICAL ENGINEERING, CBEB 2018, VOL 1, vol. 70, no. 26th Brazilian Congress on Biomedical Engineering (CBEB), R. CostaFelix, J. C. Machado, and A. V Alvarenga, Eds., Fed Univ Espirito Santo UFES, Postgrad Program Biotechnol, Vitoria, ES, Brazil, 2019, pp. 315–321. doi: 10.1007/978-981-13-2119-1_48.

[13] J. W. Choi, B. H. Kim, S. Huh, and S. Jo, “Observing Actions Through Immersive Virtual Reality Enhances Motor Imagery Training,” IEEE Transactions on Neural Systems and Rehabilitation Engineering, vol. 28, no. 7, pp. 1614–1622, Jul. 2020, doi: 10.1109/TNSRE.2020.2998123.

[14] R. Spicer, J. Anglin, D. M. Krum, and S.-L. Liew, “REINVENT: A low-cost, virtual reality brain-computer interface for severe stroke upper limb motor recovery,” in 2017 IEEE Virtual Reality (VR), Univ Southern Calif, Los Angeles, CA 90089 USA: IEEE, 2017, pp. 385–386. doi: 10.1109/VR.2017.7892338.

[15] C. A. Stefano Filho, J. Ignacio Serrano, R. Attux, G. Castellano, E. Rocon, and M. D. del Castillo, “Reorganization of Resting-State EEG Functional Connectivity Patterns in Children with Cerebral Palsy Following a Motor Imagery Virtual-Reality Intervention,” Applied Sciences, vol. 11, no. 5, p. 2372, Mar. 2021, doi: 10.3390/app11052372.

[16] A. Moldoveanu et al., “The TRAVEE System for a Multimodal Neuromotor Rehabilitation,” IEEE Access, vol. 7, pp. 8151–8171, 2019, doi: 10.1109/ACCESS.2018.2886271.

[17] R. Leeb, M. Lancelle, V. Kaiser, D. W. Fellner, and G. Pfurtscheller, “Thinking Penguin: Multimodal Brain–Computer Interface Control of a VR Game,” IEEE Trans Comput Intell AI Games, vol. 5, no. 2, pp. 117–128, Jun. 2013, doi: 10.1109/TCIAIG.2013.2242072.

[18] J. Llobera, M. González-Franco, D. Perez-Marcos, J. Valls-Solé, M. Slater, and M. V. Sanchez-Vives, “Virtual reality for assessment of patients suffering chronic pain: A case study,” Exp Brain Res, vol. 225, no. 1, pp. 105–117, Mar. 2013, doi: 10.1007/s00221-012-3352-9.

[19] D. Nath et al., “Clinical potential and neuroplastic effect of targeted virtual reality based intervention for distal upper limb in post-stroke rehabilitation: a pilot observational study,” Disabil Rehabil, vol. 0, no. 0, pp. 1–10, 2023, doi: 10.1080/09638288.2023.2228690.

